# Early detection of non-small cell lung cancer using electronic health record data

**DOI:** 10.1101/2024.10.28.24316275

**Authors:** Xiudi Li, Erin Y. Yuan, Stephen J. Kuperberg, Clara-Lea Bonzel, Mary I. Jeffway, Tianrun Cai, Katherine P. Liao, Raquel Aguiar-Ibáñez, Yu-Han Kao, Melissa L. Santorelli, David C. Christiani, Tianxi Cai, Rui Duan

## Abstract

**Rationale:** Specific patient characteristics increase the risk of cancer, necessitating personalized healthcare approaches. For high-risk individuals, tailored clinical management ensures proactive monitoring and timely interventions. Electronic Health Records (EHR) data are crucial for supporting these personalized approaches, improving cancer prevention and early diagnosis.

**Objectives:** We leverage EHR data and build a prediction model for early detection of non-small cell lung cancer (NSCLC).

**Methods:** We utilize data from Mass General Brigham’s EHR and implement a three-stage ensemble learning approach. Initially, we generate risk scores using multivariate logistic regression in a self-control and case-control design to distinguish between cases and controls. Subsequently, these risk scores are integrated and calibrated using a prospective Cox model to develop the risk prediction model.

**Results:** We identified 127 EHR-derived features predictive for early detection of NSCLC. The highly predictive features include smoking, relevant lab test results, and chronic lung diseases. The predictive model reached area under the ROC curve (AUC) of 0.801 (positive predictive value (PPV) 0.0173 with specificity 0.02) for predicting one-year NSCLC risk in a population aged 18 and above, and AUC of 0.757 (PPV 0.0196 with specificity 0.02) in a population aged 40 and above.

**Conclusions:** This study identified EHR derived features which are predictive of early NSCLC diagnosis. The developed risk prediction model exhibits superior performance for early detection of NSCLC compared to a baseline model that only relies on demographic and smoking information, demonstrating the potential of incorporating EHR derived features for personalized cancer screening recommendations and early detection.

## 1 INTRODUCTION

Cancer is the second leading cause of death in the United States, after heart disease (1). Late diagnosis is a major contributing factor to poor survival in cancer patients. According to the Surveillance, Epidemiology, and End Results (SEER) program of the National Cancer Institute, between 2014 and 2020, over half of lung and pancreatic cancers and nearly a quarter of colorectal cancer are being diagnosed at a metastatic stage when treatment is less effective and overall prognosis is poor (2). Cancer detected early at stage I has a much higher five-year survival rate than if it is detected later (3). Besides the difference in overall survival, patients with cancer who are diagnosed earlier are also more likely to have lower treatment morbidity, better experience of care and improved quality of life than those diagnosed late (4). While cancers such as breast cancer have easily identifiable early signs, cancers in the lung usually lack early symptoms (5, 6), leading to delayed detection. Also, some cancer types including colorectal and breast cancer have well-developed screening guidelines, while lung cancer screening is only recommended for high-risk individuals identified based on age and smoking history (7, 8) and the uptake can be low (9). Therefore, there is a need to employ methods for diagnosis of lung cancer at an earlier stage, enabling early treatment which leads to improved prognosis and survival.

The US Preventive Services Task Force (USPSTF) recommends annual screening for lung cancer with low-dose CT (LDCT) in adults aged 50 to 80 years with a 20-pack-year smoking history and currently smoking or who have quit smoking within the past 15 years (8). Also, the USPSTF concluded that lung cancer screening with LDCT has a moderate net benefit in persons at high risk of lung cancer (8). However, the national uptake of lung cancer screening was estimated at <6% among USPSTF-criteria eligible smokers in 2015 (9). Moreover, important patient subpopulations such as younger patients and non-smokers are not eligible for screening under the current guidelines. Therefore, developing risk prediction tools and screening programs leveraging routinely collected healthcare visit data has huge potential in complementing the current screening guidelines and improving early detection of lung cancer.

The estimated number of new lung cancer cases in the US in 2024 is 234,580, and the estimated number of lung cancer deaths is 125,070 (10). As per SEER, the 5-year survival rate for lung cancer in the US between 2014 and 2020 increased to 26.7% compared to 11.5% in 1975 (2, 11). This increase in survival rate is likely due to improved targeted treatment and immunotherapies. The primary factor affecting lung cancer prognosis is the stage at diagnosis, and lung cancer is usually diagnosed at a late stage contributing to poor prognosis and low overall survival rate (12). In particular, between 2014 and 2020, 22% of lung cancer cases are diagnosed at stage I-II, 21% at stage III, and 53% at stage IV (1); the 5-year survival rate for patients with localized disease (i.e., stages I-II) at diagnosis is 63.7%, while that for patients with regional disease (stage III) is 35.9%, and 8.9% for patients with metastatic disease (stage IV) (2).

Clinical prediction models are increasingly valuable in providing healthcare professionals and patients with insights into potential health outcomes, thereby enhancing medical decision-making and improving overall health outcomes. Electronic Health Records (EHR) data, collected through routine healthcare visits, serve as a rich source for training these models. This data enables the seamless integration of clinical risk prediction models into health systems, further optimizing patient care. Previous studies have developed risk prediction models for different cancer types including lung cancer. One of the most well-known lung cancer risk prediction models is the PLCO^m^2012 model (13–15), but it relies on detailed information on smoking which may not be well-captured in the EHR and does not fully utilize the rich patient history in the EHR. In a study conducted by Muller et.al., various predictors of lung cancer risk and lung function (including functional expiratory volume, FEV1) were included in a prediction model to assess the predictive ability of lung function in a population-based cohort to improve model performance for lung cancer risk prediction (16). However, measures of lung function may not be routinely available in the EHR. In another study conducted by Gould et.al., a machine learning model was proved to be more accurate for early diagnosis of non-small cell lung cancer (NSCLC) than standard screening eligibility criteria based on age and smoking history, showing potential to prevent lung cancer deaths through early detection (17). However, the model was trained using a case-control design and was not evaluated prospectively to predict incident lung cancer. In another study that developed and validated a prospective risk prediction model for identifying patients at risk of new incident lung cancer, researchers utilized EHR data to construct an algorithm that achieved an accuracy of 88% (18). It is important to note that this study did not focus on early-stage lung cancer and included previous diagnoses of other cancers as predictors. This introduces the possibility that the lung cancer may be secondary, and limits the applicability of the model, as patients with prior cancer diagnosis would typically be under specialized care and monitoring. A recent study by Chandran et al (19) built a prediction model for all lung cancer diagnosis and did not focus on early-stage diagnosis either. Furthermore, their model only utilized the codified data but not the data from clinical notes.

In this paper, we develop and prospectively evaluate a risk prediction model for the early diagnosis of NSCLC using routine healthcare data. This model aims to identify patients at high risk of NSCLC, enabling regular monitoring and screening to increase the likelihood of detecting lung cancer at an early stage. Additionally, the model may also be used to help with early diagnosis among NSCLC patients who may be undiagnosed, facilitating earlier referrals to specialized care and treatment to improve prognosis and survival. While the primary focus is on early-stage detection, we also developed and evaluated risk prediction models for all stages of NSCLC to compare the differences in key features between stages, which helps in understanding by identifying which features are most predictive in early stages compared to later ones, potentially enhancing early prevention of NSCLC.

## 2 METHODS

### 2.1 Study Design

This study used two EHR cohorts extracted from the Mass General Brigham (MGB) EHR databases. The first cohort, referred to as the Lung Cancer (LC) data mart, represented a cohort of lung cancer patients, comprising individuals identified (retrospectively) with lung cancer using an established machine learning algorithm (20). The second is the MGB Biobank cohort (21), which enrolled individuals from the broader MGB EHR population. Codified features were extracted from structured data, including demographics, diagnoses, medications, labs, and procedures. We also curated narrative features extracted via natural language processing (NLP) from narrative clinical texts to infer cancer-related information such as lifestyle (e.g., smoking status, alcohol consumption), and other relevant health conditions (22). All NLP concepts were standardized to concept unique identifiers (CUI) according to the Unified Medical Language System (UMLS) (23). Histologic type and tumor stage information were inferred from clinical notes using an established machine learning algorithm (20). Additionally, features related to socioeconomic status, including social vulnerability index (24) and area deprivation index on the national and state level (25), were derived from five-digit zip codes. Leveraging the two cohorts, this study used the three-stage Prospective-retrospective hybrid design (POETRY) developed by Li et al (26), which is illustrated in Figure 1.

**Figure 1:**
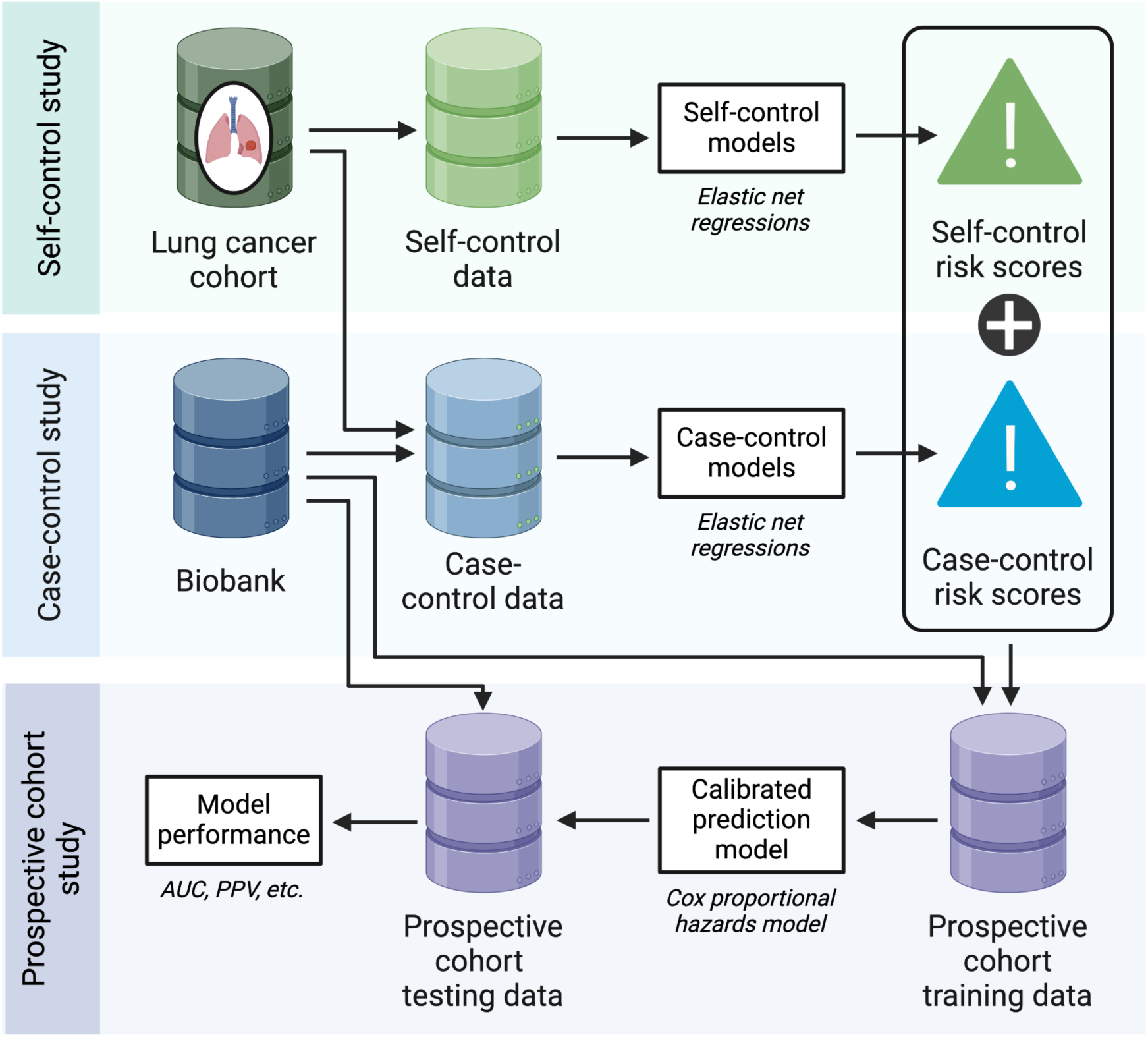
Overview of the study design

**Figure 2:**
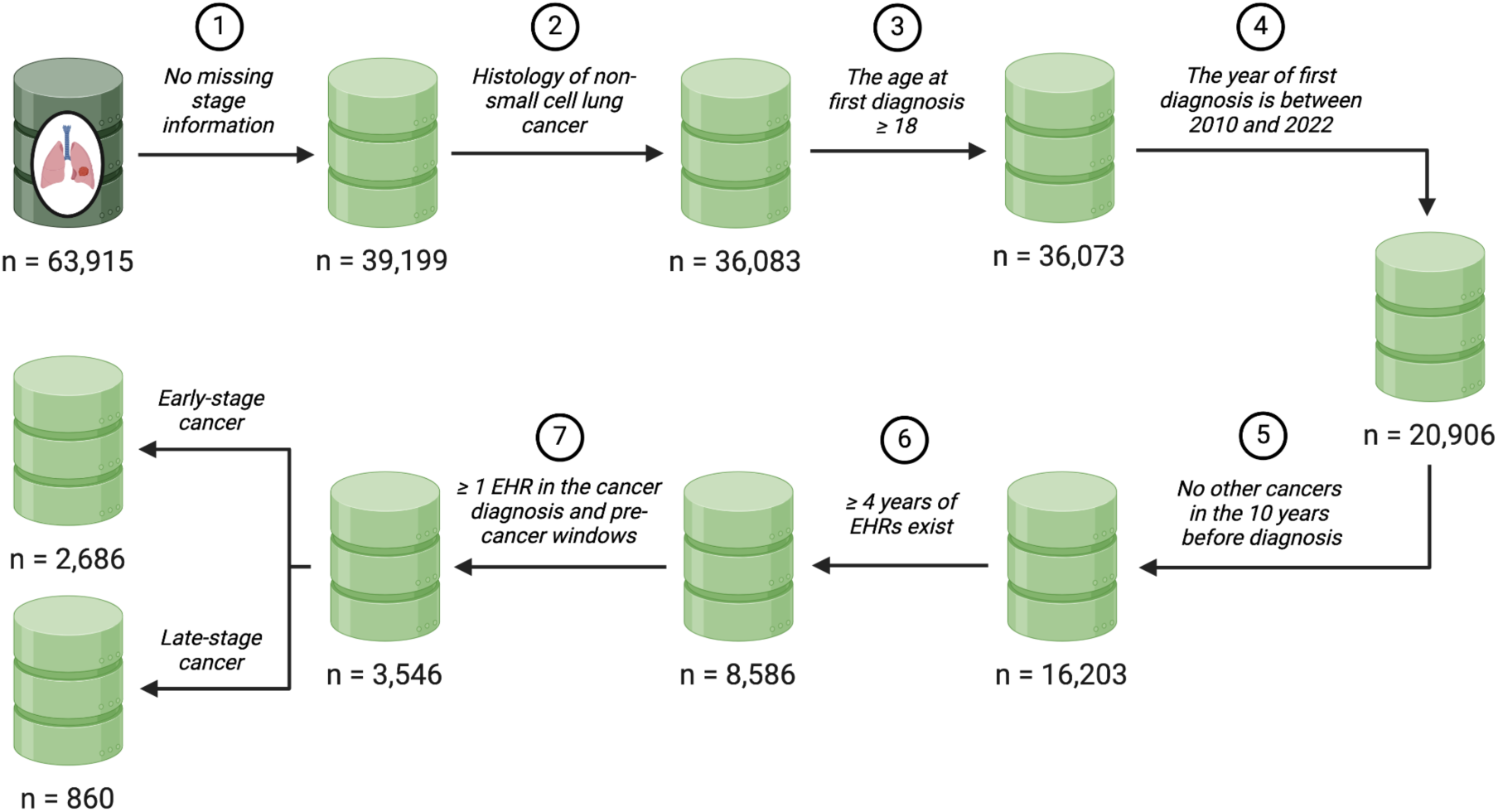
Construction of the study sample in the self-control design

As illustrated in Figure 1, the study employs the POETRY approach (26) consisting of three key components: (i) a self-control design comparing data from pre-cancer window to a post-cancer window of LC cases; (ii) a case-control design comparing LC cases to matched controls; and (iii) a prospective design that combines risk scores derived from the self-control and case-control designs to form a final prediction model.

Using a self-control design, we formed an all-case cohort from the LC Mart, detailed in Section 2.2 with specific inclusion and exclusion criteria. We defined two longitudinal time windows around the first NSCLC diagnosis date: the cancer diagnosis window which is the one-year window prior to the date of the first lung cancer diagnosis, and the preceding one-year pre-cancer control window at least two years ahead of the cancer diagnosis window. By comparing these periods as "digital twins," we controlled for confounding effects and identified early, relevant features that changed over time.

The case-control design was also conducted to help identify features that were less time-varying. For a case in the NSCLC cohort, we matched it to two lung-cancer-free patients from the MGB Biobank, and matching was based on patient demographics including age, sex, and race, healthcare utilization, and the calendar year of the control window. By comparing cases with lung-cancer-free controls, features related to cancer diagnosis were identified.

In both the self-control and case-control designs, not only were features associated with lung cancer diagnosis identified, but classification models were also developed to generate risk scores. These risk scores helped distinguish between cases and controls, as well as the cancer diagnosis period from the pre-cancer period.

In the subsequent prospective modeling stage, we develop a risk prediction model by integrating risk scores from classification models trained using self-control and case-control designs, utilizing data from the MGB Biobank. For each calendar year from 2007 to 2021, we defined a cohort with a one-year feature window and set the outcome as the time to lung cancer diagnosis one year after the feature window. An ensemble learning technique was applied to integrate and calibrate the risk scores from both designs effectively.

### 2.2 Study Population

To form an NSCLC all-case cohort, patients were selected from the LC Mart based on several criteria: diagnosis of lung cancer between January 2006 and December 2021, confirmed NSCLC histologic type and cancer stage, age of 18 or older at first diagnosis, diagnosis after 2010 for the self-control design and after 2007 for the case-control design^1^, at least four years of observation prior to diagnosis, no prior cancer diagnosis for ten years, and EHR records availability during both diagnosis and pre-diagnosis periods. Patients were further categorized into early-stage (Stage I-III) or metastatic NSCLC cohorts based on their cancer stage. Additionally, the MGB Biobank was used for a case-control design, matching each NSCLC patient with two controls free from lung cancer, based on year, demographics, and healthcare utilization. Yearly cohorts from 2007 to 2021 were also constructed using the MGB Biobank, with inclusion criteria including enrollment in the Biobank, EHR records from at least one year prior to January 1^st^ of the cohort year, no lung cancer history or other cancer diagnoses in the ten years before January 1^st^ of cohort year.

### 2.3 Feature and Outcome Curation

To curate EHR features and outcome for modeling, all EHR codified data were standardized and normalized into higher level concepts, which are more clinically meaningful compared to detailed EHR codes, according to existing ontologies. Diagnostic codes were grouped into PheCodes according to the PheWAS catalog (27). All procedure codes were grouped into procedure groups such as “MRI” according to the Clinical Classification System (28). Medication codes were grouped into RxNorm (29), and laboratory test codes into Logical Observation Identifiers Names and Codes (LOINC) (30). Narrative data is processed through NLP tools like NILE (22), extracting mentions of clinical terms as Concept Unique Identifiers (CUIs).

An initial list of codified features was generated using the Online Narrative and Codified feature Search Engine (ONCE), which creates a knowledge graph along with semantic embedding vectors for a large number of EHR concepts including both codified concepts such as PheCodes and RxNorm and NLP concepts covering many semantic types such as disorders and laboratory procedures (31). The cosine similarity between two embedding vectors associated with two concepts quantifies the degree of relatedness between the two concepts. We selected codified and NLP concepts that have cosine similarity higher than 0.2 with the target disease PheCode:165.1(lung cancer). We additionally augmented the NLP concept list with domain expert input. A complete list can be found in the online supplementary material (see Tables A7 and A8). All selected codified or NLP concepts were extracted across the entire follow up time window for each patient.

For each design, we created predictive features to represent each concept from the corresponding time windows, which are defined as the log total counts of each concept within the window. Additionally, a measure of healthcare utilization was computed as the log total count of all integer-level phecodes (diagnosis codes). Binary outcome was defined, taking value 1 for cases in the case-control design (or cancer diagnosis window in the self-control design) and 0 for controls (or the pre-cancer control window). For the third stage of prospective analysis, we split the MGB Biobank patient subset not used in the case-control study into two groups: one for training the calibration model (calibration fold) and the other for performance evaluation (evaluation fold). We established yearly cohorts from 2007 to 2021, allowing patients to appear in multiple cohorts if they met the inclusion criteria (refer to Section 2.2).

For each year, we utilized the prior year as the feature window to compute the log count of EHR features and healthcare utilization. The time to NSCLC diagnosis within the year served as the outcome, which was censored at year’s end if no lung cancer was present or due to follow-up loss or small cell lung cancer diagnosis. We then compiled a stacked calibration dataset from the annual data of patients in the calibration fold and similarly constructed an evaluation dataset using the evaluation fold data.

## 3 STATISTICAL ANALYSES

### Feature Screening

We first conducted feature screening to identify informative features for further modeling using both self-control design and case-control design. Feature frequencies were calculated for cases and controls separately, defined as the proportion of patients having a non-zero count, and only features with frequency at least 1% in at least one of the windows were kept. Further screening of each feature based on marginal association was then conducted. Specifically, we fitted a conditional logistic regression in the self-control design including one feature at a time as the covariate, adjusting for healthcare utilization, and stratified by patient. In the case-control design, we employed logistic regression including one EHR feature at a time as the covariate, adjusting for all variables used in the matching. Marginal screening based on p-values was conducted for both designs, and the false discovery rate was controlled at 0.05 via the Benjamini-Hochberg approach (32). Finally, from all the features that pass the screening, we excluded features that were directly associated with lung cancer diagnosis such as CT scans and nodule findings.

For lab tests in LOINC codes which passed the marginal screening, we extracted the numerical values from the EHR and included them as additional features. To handle the missingness, a last-observation-carry-forward approach was applied, where the missing lab value in the cancer diagnosis/pre-cancer control window was replaced with the last observed value within two years prior to the start of the window. If no lab value was observed within the two years prior, the lab value was left missing. The remaining missing values were imputed using Multivariate Imputation by Chained Equations (MICE), including patient demographics and numeric lab values in the imputation model (33).

### Model Development

For both the self-control design and case-control design, classification models were built via multivariate logistic regression. Patient demographics (age, gender, and race), imputed lab values, lab value missing indicators, log counts of EHR features (codes and CUIs) that passed the marginal screening in both the self-control design and the case-control design, and healthcare utilization were used as predictors, and an elastic net penalty was applied. Five different tuning parameter values for the elastic net penalty were used to interpolate between ridge regression and lasso regression, encouraging different levels of sparsity in the regression model. The penalty parameter was selected via cross-validation. This resulted in a set of 5 classification models in each design, leading to 10 risk scores for each patient at any given time window.

In the third stage, a Cox proportional hazard model was fitted to the calendar-time specific prospective cohorts as described above using the 10 risk scores along with patient demographics and healthcare utilization as predictors, stratified by 3-year periods to allow for changes in the baseline hazard over calendar time, reflecting temporal shifts in lung cancer risk. Additionally, we developed a baseline model using only patient demographics and smoking information for comparison. The predictive performance was measured using the area under the receiver operating characteristics curve (ROC) and positive predictive value (PPV), both overall and by 3-year periods, calculated on the evaluation dataset.

For our main analysis, we trained all models using early-stage NSCLC patients aged 18 and above. As a sensitivity analysis, two additional populations were considered: (1) including all (early- and late-stage) NSCLC patients; (2) only focusing on patients aged 40 and above at diagnosis. Our focus is on populations younger than current guidelines by USPSTF, as early detection for younger people at risk may increase the overall years of life saved (34).

## 4 RESULTS

### 4.1 Construction of Study Sample

#### 4.1.1 Self-control design

Among the initial 63,915 patients in the LC Mart, we selected 3,546 eligible patients, among which 2,686 were diagnosed at an early stage (stage I to III) at the time of their first lung cancer diagnosis, while 860 patients were diagnosed at a metastatic stage. Table 1 presents the relevant patient characteristics for these eligible patients. Furthermore, for a comprehensive overview of patient characteristics for the entire lung cancer cohort, please refer to Table A1 in the Supplementary Material.

**Table 1:**
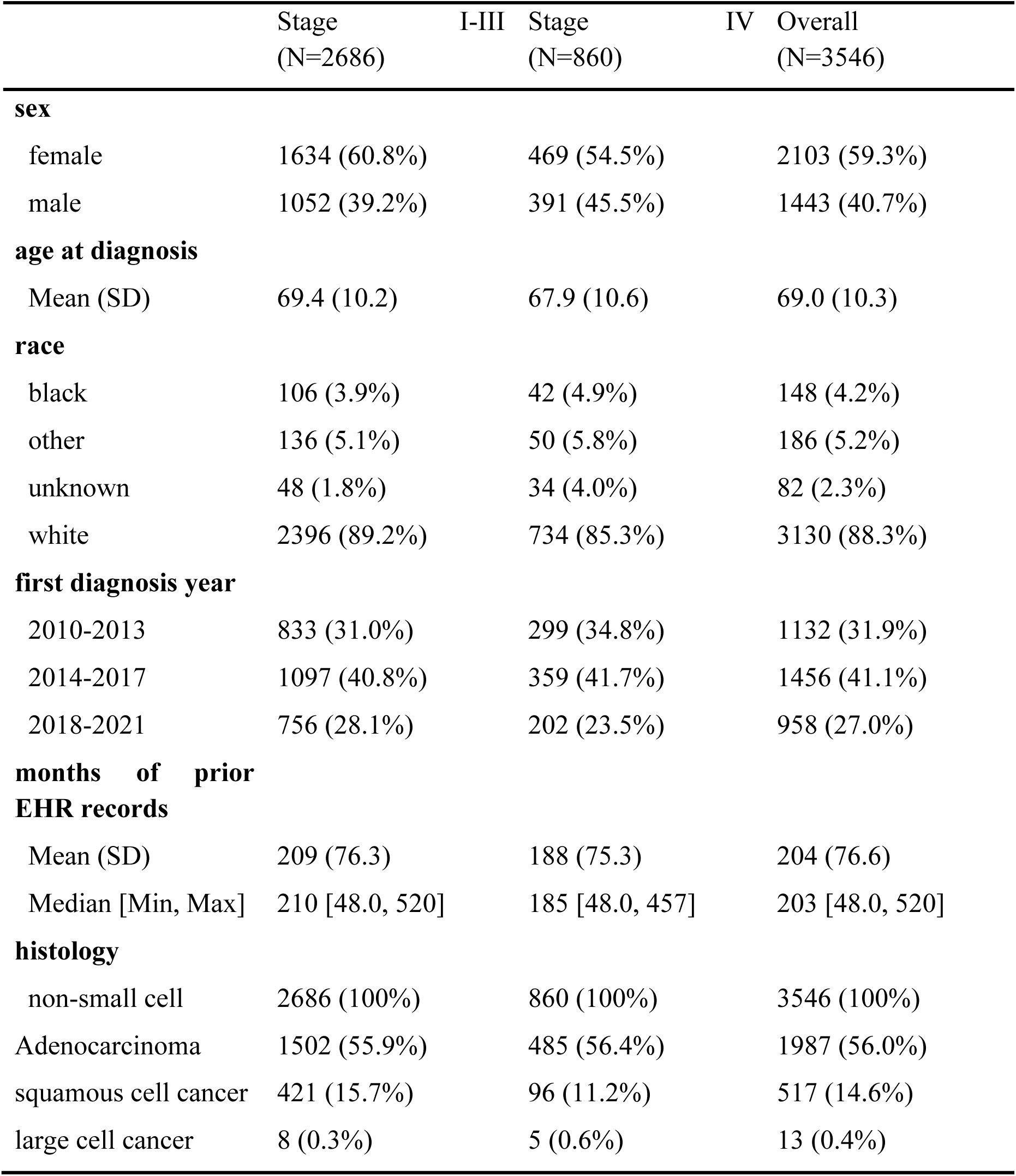
Demographics and clinical attributes of the patients in the lung cancer cohort that are eligible for the self-control analysis, overall and stratified by stage (stage I-III versus stage IV). For continuous variables we present mean (standard deviation), and for categorical variables we present count (percentage).

When using a self-control design, time-invariant patient demographics, such as race and sex, remain constant during both the cancer diagnosis period and the pre-cancer control period. In

Table 2, we provided a summary of smoking status, comorbidities, and lab information that may change over time for eligible early-stage patients. A similar table was presented in the Supplementary Material for all eligible patients (see Table A2). Three lab test results with a high missing rate (>95%) are excluded from the analysis.

**Table 2:**
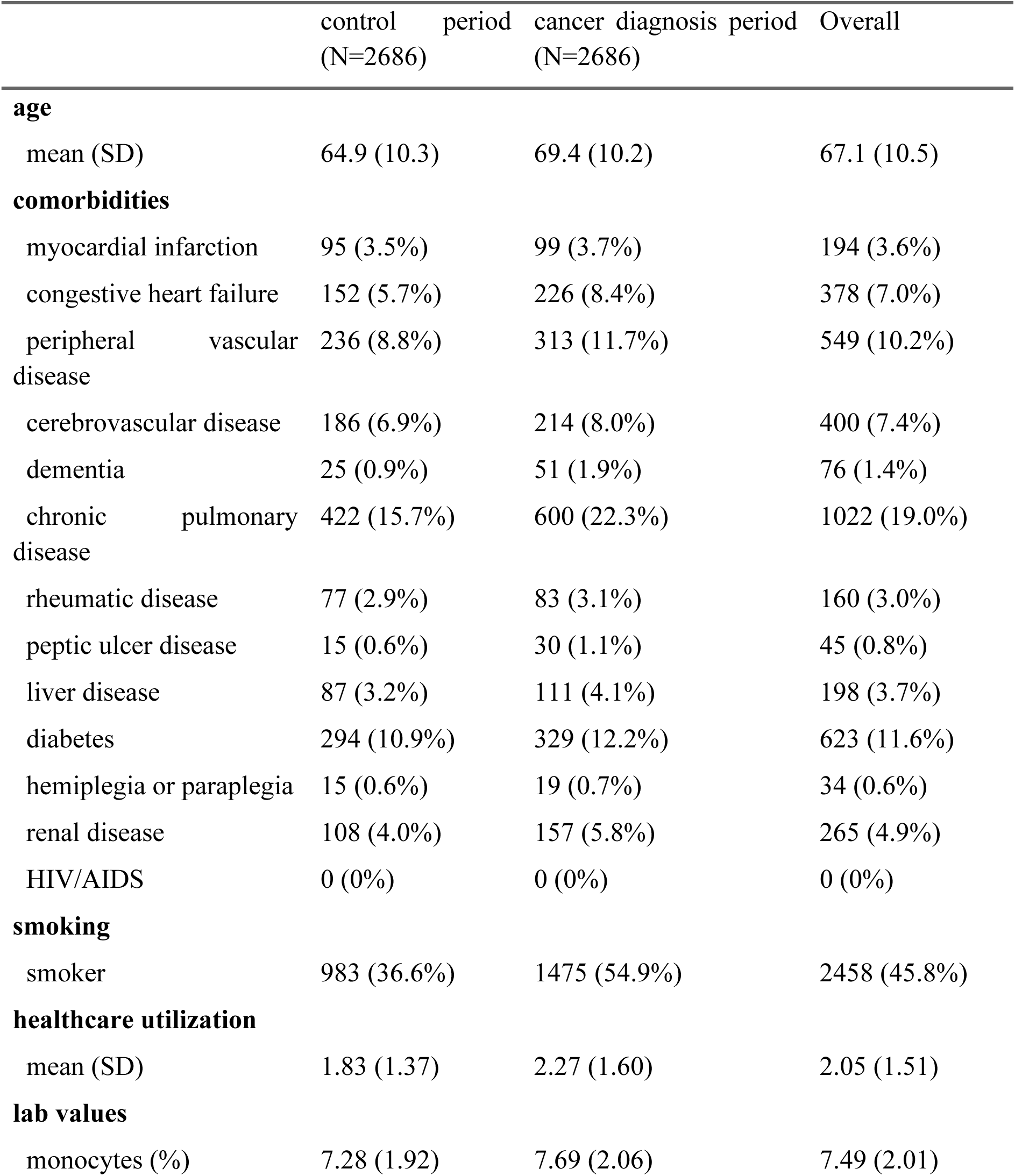
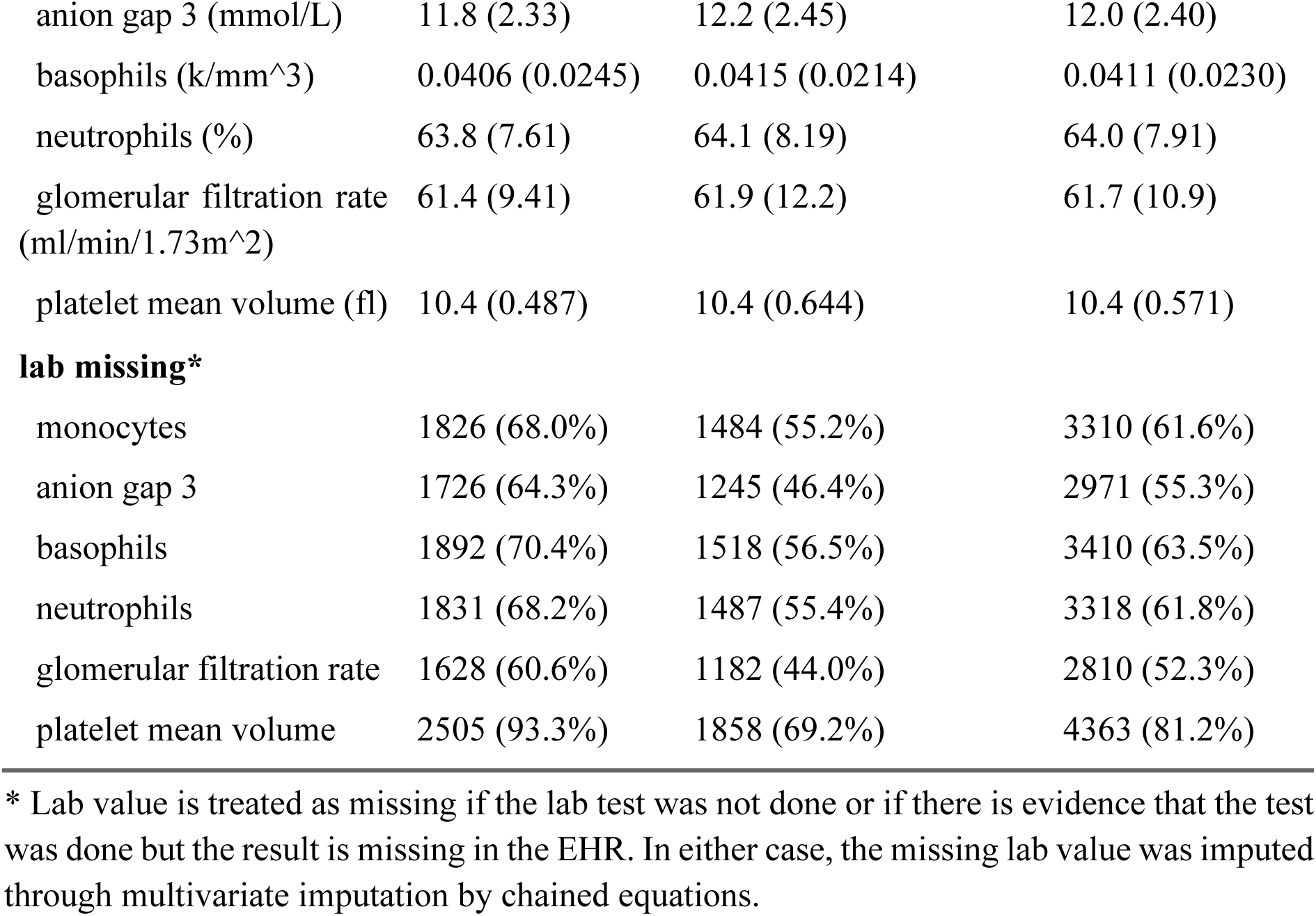
Age, smoking, comorbidities, and lab values comparing cancer-diagnosis and control windows among early-stage NSCLC patients in the lung cancer cohort who met all inclusion criteria. Smoking is defined as the occurrence of smoking-related code and/or CUI; comorbidity is defined as the occurrence of relevant diagnosis codes.

#### 4.1.2 Case-control design

There are 6,354 NSCLC eligible patients who were included in the case-control study. They were matched to 12,686 controls from the MGB Biobank, who had no history of lung cancer. In Table 3, we provide a comprehensive summary of the baseline characteristics of eligible early-stage patients and their matched controls. Additionally, a similar table on all NSCLC patients and their matched controls is available in the Supplementary Material (see Table A3.)

**Table 3:**
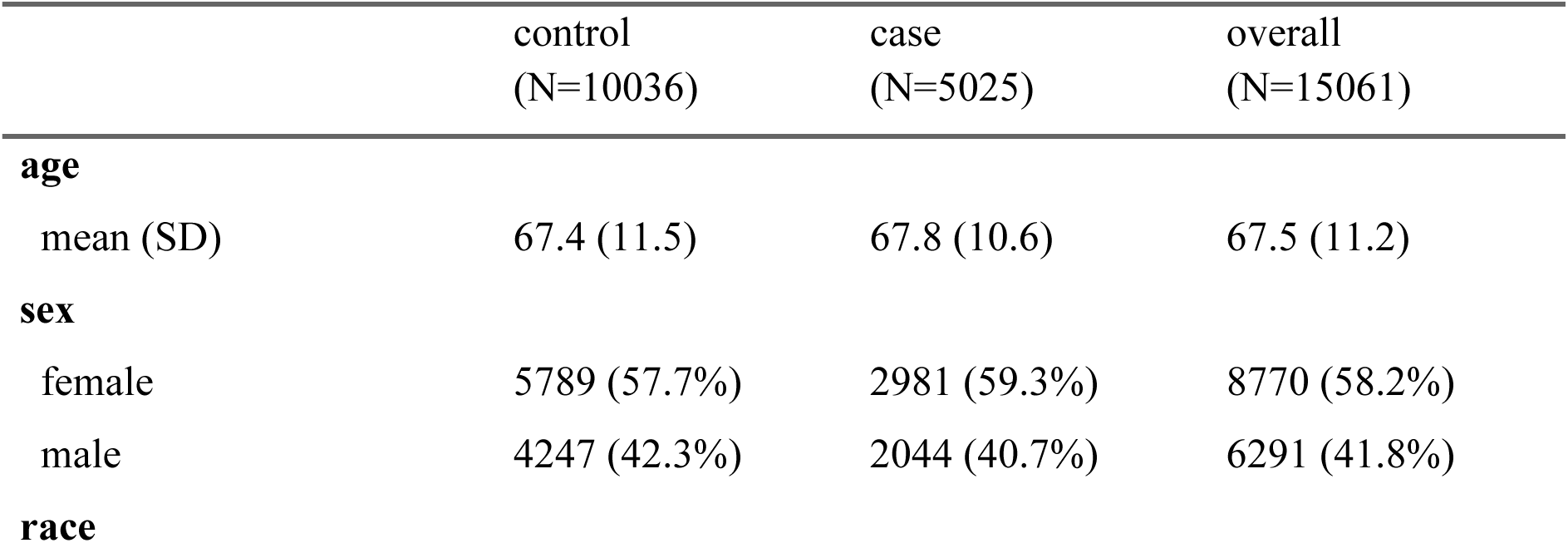
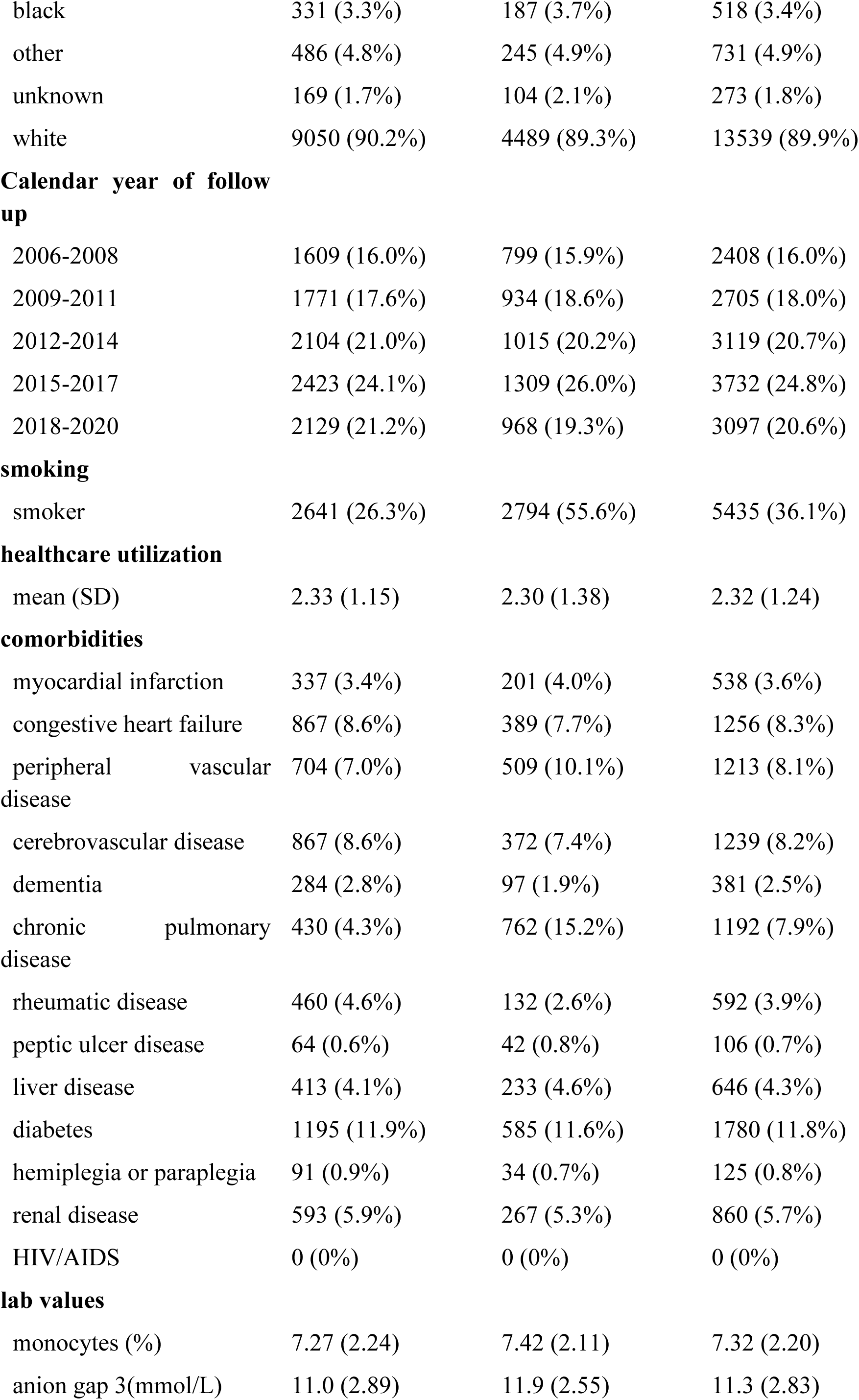
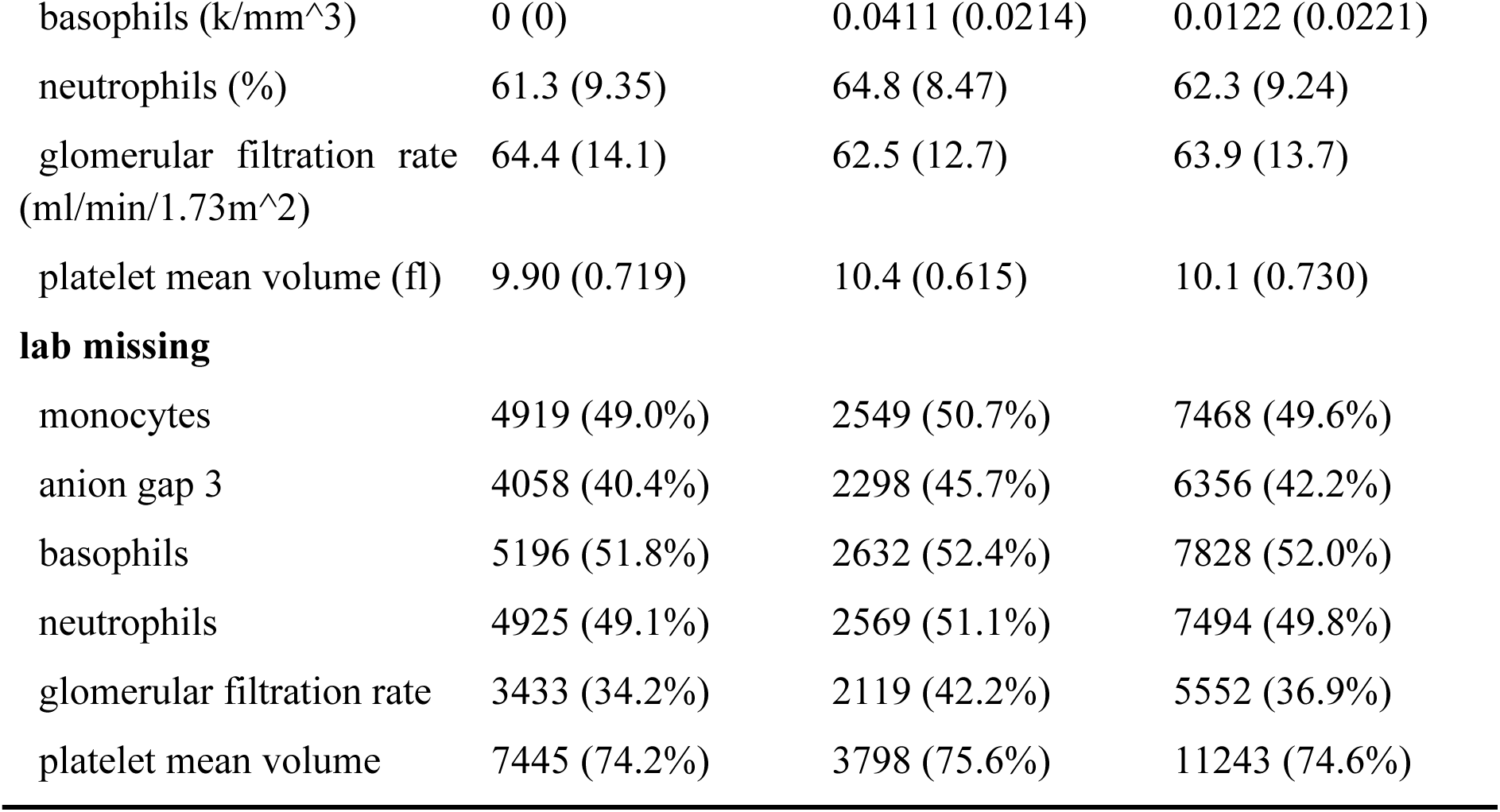
Baseline characteristics of patients in the case-control design. Cases are early-stage NSCLC patients from the lung cancer cohort; and controls are patients free of lung cancer in the MBG Biobank, matched based on calendar year, healthcare utilization, and patient demographics.

|  | control<br>(N=10036) | case<br>(N=5025) | overall<br>(N=15061) |
| --- | --- | --- | --- |
| **age** |  |  |  |
| mean (SD) | 67.4 (11.5) | 67.8 (10.6) | 67.5 (11.2) |
| **sex** |  |  |  |
| female | 5789 (57.7%) | 2981 (59.3%) | 8770 (58.2%) |
| male | 4247 (42.3%) | 2044 (40.7%) | 6291 (41.8%) |
| **race** |  |  |  |
| black | 331 (3.3%) | 187 (3.7%) | 518 (3.4%) |
| other | 486 (4.8%) | 245 (4.9%) | 731 (4.9%) |
| unknown | 169 (1.7%) | 104 (2.1%) | 273 (1.8%) |
| white | 9050 (90.2%) | 4489 (89.3%) | 13539 (89.9%) |
| **Calendar year of follow up** |  |  |  |
| 2006-2008 | 1609 (16.0%) | 799 (15.9%) | 2408 (16.0%) |
| 2009-2011 | 1771 (17.6%) | 934 (18.6%) | 2705 (18.0%) |
| 2012-2014 | 2104 (21.0%) | 1015 (20.2%) | 3119 (20.7%) |
| 2015-2017 | 2423 (24.1%) | 1309 (26.0%) | 3732 (24.8%) |
| 2018-2020 | 2129 (21.2%) | 968 (19.3%) | 3097 (20.6%) |
| **smoking** |  |  |  |
| smoker | 2641 (26.3%) | 2794 (55.6%) | 5435 (36.1%) |
| **healthcare utilization** |  |  |  |
| mean (SD) | 2.33 (1.15) | 2.30 (1.38) | 2.32 (1.24) |
| **comorbidities** |  |  |  |
| myocardial infarction | 337 (3.4%) | 201 (4.0%) | 538 (3.6%) |
| congestive heart failure | 867 (8.6%) | 389 (7.7%) | 1256 (8.3%) |
| peripheral vascular disease | 704 (7.0%) | 509 (10.1%) | 1213 (8.1%) |
| cerebrovascular disease | 867 (8.6%) | 372 (7.4%) | 1239 (8.2%) |
| dementia | 284 (2.8%) | 97 (1.9%) | 381 (2.5%) |
| chronic pulmonary disease | 430 (4.3%) | 762 (15.2%) | 1192 (7.9%) |
| rheumatic disease | 460 (4.6%) | 132 (2.6%) | 592 (3.9%) |
| peptic ulcer disease | 64 (0.6%) | 42 (0.8%) | 106 (0.7%) |
| liver disease | 413 (4.1%) | 233 (4.6%) | 646 (4.3%) |
| diabetes | 1195 (11.9%) | 585 (11.6%) | 1780 (11.8%) |
| hemiplegia or paraplegia | 91 (0.9%) | 34 (0.7%) | 125 (0.8%) |
| renal disease | 593 (5.9%) | 267 (5.3%) | 860 (5.7%) |
| HIV/AIDS | 0 (0%) | 0 (0%) | 0 (0%) |
| **lab values** |  |  |  |
| monocytes (%) | 7.27 (2.24) | 7.42 (2.11) | 7.32 (2.20) |
| anion gap 3(mmol/L) | 11.0 (2.89) | 11.9 (2.55) | 11.3 (2.83) |
| basophils (k/mm<sup>3</sup>) | 0 (0) | 0.0411 (0.0214) | 0.0122 (0.0221) |
| neutrophils (%) | 61.3 (9.35) | 64.8 (8.47) | 62.3 (9.24) |
| glomerular filtration rate (ml/min/1.73m<sup>2</sup>) | 64.4 (14.1) | 62.5 (12.7) | 63.9 (13.7) |
| platelet mean volume (fl) | 9.90 (0.719) | 10.4 (0.615) | 10.1 (0.730) |
| **lab missing** |  |  |  |
| monocytes | 4919 (49.0%) | 2549 (50.7%) | 7468 (49.6%) |
| anion gap 3 | 4058 (40.4%) | 2298 (45.7%) | 6356 (42.2%) |
| basophils | 5196 (51.8%) | 2632 (52.4%) | 7828 (52.0%) |
| neutrophils | 4925 (49.1%) | 2569 (51.1%) | 7494 (49.8%) |
| glomerular filtration rate | 3433 (34.2%) | 2119 (42.2%) | 5552 (36.9%) |
| platelet mean volume | 7445 (74.2%) | 3798 (75.6%) | 11243 (74.6%) |

**4.1.3 Prospective risk modeling**

Figure 3 illustrates the selection of patients to construct the yearly cohorts for prospective model fitting. In Table 4, we provide the total sample sizes and the number of NSCLC and early-stage NSCLC cases in each yearly cohort. After stacking the data from all yearly cohorts, we obtained 56,3817 observations where each observation corresponds to a patient-year. The overall incidence rate of NSCLC is 0.21%, and 0.16% for early-stage diagnosis.

**Figure 3:**
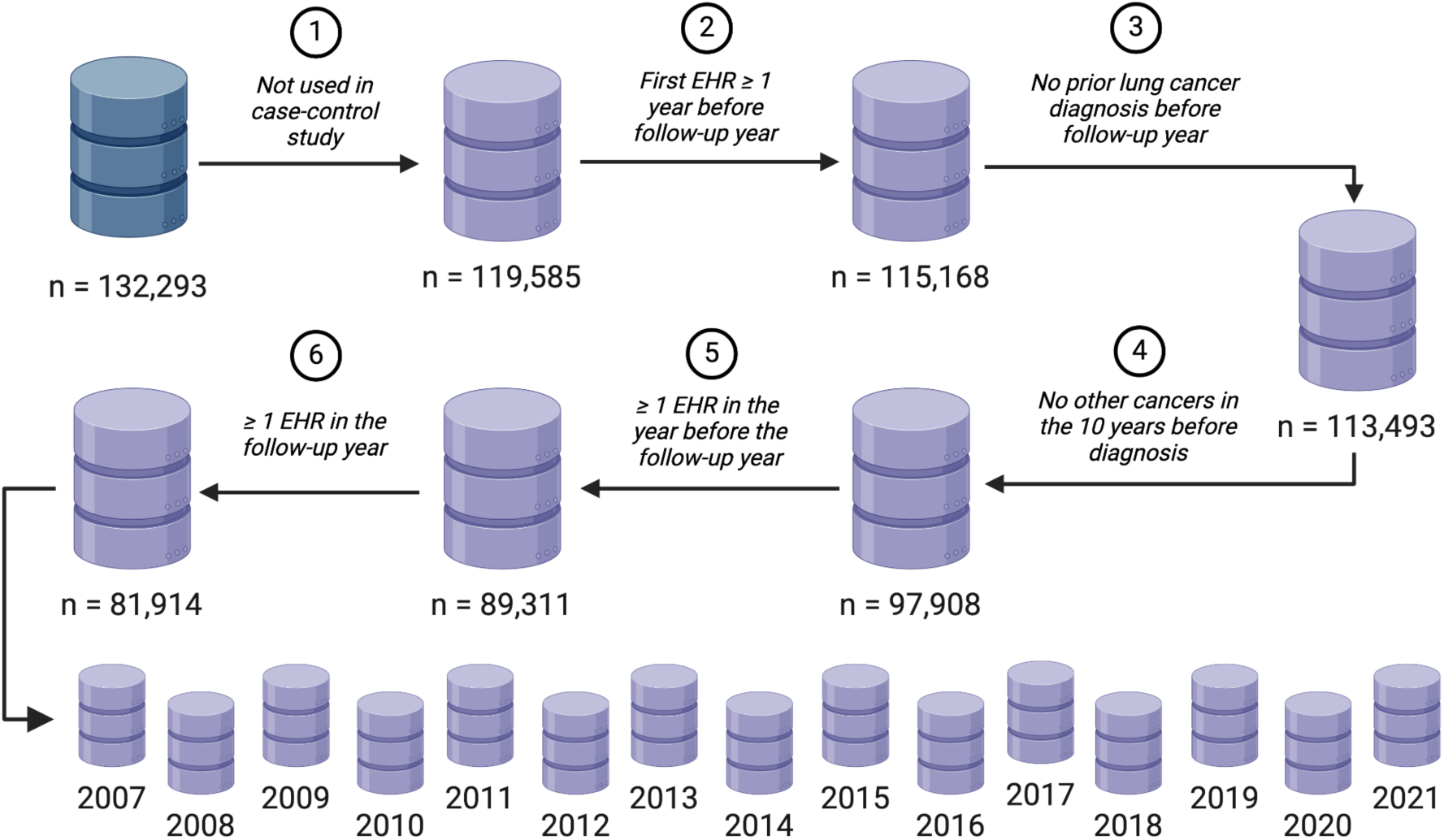
Construction of the study sample in prospective modeling

**Table 4:** Number of eligible patients and number of NSCLC cases in yearly cohorts in the cohort data from MGB Biobank.

|  | 2007 | 2008 | 2009 | 2010 | 2011 | 2012 | 2013 | 2014 |
| --- | --- | --- | --- | --- | --- | --- | --- | --- |
| **eligible patients** | 23192 | 24774 | 26704 | 28425 | 30181 | 32496 | 34994 | 37343 |
| **NSCLC cases** | 25 | 27 | 33 | 44 | 48 | 46 | 76 | 69 |
| **early-stage**<br>**NSCLC cases** | 25 | 22 | 32 | 43 | 40 | 40 | 61 | 53 |
|  | 2015 | 2016 | 2017 | 2018 | 2019 | 2020 | 2021 | total |
| **eligible patients** | 40187 | 43336 | 45875 | 48079 | 50483 | 51032 | 46716 | 563817 |
| **NSCLC cases** | 103 | 125 | 127 | 167 | 163 | 101 | 26 | 1180 |
| **early-stage**<br>**NSCLC cases** | 88 | 90 | 86 | 117 | 102 | 71 | 19 | 889 |

We summarize patient demographics, smoking status, comorbidities, and lab tests in Table 5, comparing early-stage NSCLC cases versus NSCLC-free patients. In the Supplementary Material, we give the same comparison on all NSCLC cases (see Table A4.)

**Table 5:** Patient demographics, smoking status, comorbidities, and lab tests in the MGB Biobank patients in the prospective modeling stage, comparing early-stage NSCLC cases to lung-cancer-free patients.

|  | NSCLC-free<br>(N=562637) | early-stage NSCLC case<br>(N=889) | Overall<br>(N=563817) |
| --- | --- | --- | --- |
| **age** |  |  |  |
| mean (SD) | 50.4 (15.3) | 64.0 (11.9) | 50.4 (15.3) |
| **race** |  |  |  |
| black | 32865 (5.8%) | 30 (3.4%) | 32895 (5.8%) |
| other | 40900 (7.3%) | 30 (3.4%) | 40930 (7.3%) |
| unknown | 14674 (2.6%) | 23 (2.6%) | 14697 (2.6%) |
| white | 474198 (84.3%) | 806 (90.7%) | 475004 (84.3%) |
| **sex** |  |  |  |
| female | 326615 (58.1%) | 473 (53.2%) | 327088 (58.0%) |
| male | 236006 (41.9%) | 416 (46.8%) | 236422 (42.0%) |
| unknown | 16 (0.0%) | 0 (0%) | 16 (0.0%) |
| **area deprivation index (national)** |  |  |  |
| mean (SD) | 13.2 (12.6) | 14.8 (14.9) | 13.2 (12.6) |
| **area deprivation index (state)** |  |  |  |
| mean (SD) | 3.73 (2.36) | 4.02 (2.52) | 3.73 (2.36) |
| **social vulnerability index** |  |  |  |
| mean (SD) | 0.375 (0.198) | 0.376 (0.201) | 0.375 (0.198) |
| **calendar year** |  |  |  |
| 2007-2009 | 74585 (13.3%) | 79 (8.9%) | 74664 (13.2%) |
| 2010-2012 | 90964 (16.2%) | 123 (13.8%) | 91087 (16.2%) |
| 2013-2015 | 112276 (20.0%) | 202 (22.7%) | 112478 (20.0%) |
| 2016-2018 | 136871 (24.3%) | 293 (33.0%) | 137164 (24.3%) |
| 2019-2021 | 147941 (26.3%) | 192 (21.6%) | 148133 (26.3%) |
| **healthcare utilization** |  |  |  |
| mean (SD) | 2.55 (1.23) | 2.87 (1.43) | 2.55 (1.23) |
| **comorbidities** |  |  |  |
| myocardial infarction | 11932 (2.1%) | 38 (4.3%) | 11970 (2.1%) |
| congestive heart failure | 33657 (6.0%) | 100 (11.2%) | 33757 (6.0%) |
| peripheral vascular disease | 25806 (4.6%) | 86 (9.7%) | 25892 (4.6%) |
| cerebrovascular disease | 25027 (4.4%) | 71 (8.0%) | 25098 (4.5%) |
| dementia | 6654 (1.2%) | 20 (2.2%) | 6674 (1.2%) |
| chronic pulmonary disease | 63914 (11.4%) | 260 (29.2%) | 64174 (11.4%) |
| rheumatic disease | 26905 (4.8%) | 54 (6.1%) | 26959 (4.8%) |
| peptic ulcer disease | 3217 (0.6%) | 7 (0.8%) | 3224 (0.6%) |
| liver disease | 36806 (6.5%) | 83 (9.3%) | 36889 (6.5%) |
| diabetes | 64686 (11.5%) | 161 (18.1%) | 64847 (11.5%) |
| hemiplegia or paraplegia | 4473 (0.8%) | 11 (1.2%) | 4484 (0.8%) |
| renal disease | 27927 (5.0%) | 97 (10.9%) | 28024 (5.0%) |
| HIV/AIDS | 2730 (0.5%) | 4 (0.5%) | 2734 (0.5%) |
| **smoking** |  |  |  |
| smoker | 158453 (28.2%) | 457 (51.4%) | 158910 (28.2%) |
| **lab value** |  |  |  |
| monocytes (%) | 7.37 (2.22) | 7.72 (2.53) | 7.37 (2.22) |
| anion gap 3(mmol/L) | 11.6 (2.86) | 11.8 (2.90) | 11.6 (2.86) |
| basophils (k/mm<sup>3</sup>) | 0.0410 (0.0262) | 0.0444 (0.0267) | 0.0410 (0.0262) |
| neutrophils (%) | 61.3 (9.69) | 63.3 (9.86) | 61.3 (9.69) |
| glomerular filtration rate (ml/min/1.73m<sup>2</sup>) | 70.3 (18.0) | 64.2 (17.4) | 70.3 (18.0) |
| platelet mean volume (fl) | 10.4 (0.699) | 10.4 (0.743) | 10.4 (0.699) |
| **lab missing** |  |  |  |
| monocytes | 239650 (42.6%) | 332 (37.3%) | 239982 (42.6%) |
| anion gap 3 | 200981 (35.7%) | 294 (33.1%) | 201275 (35.7%) |
| basophils | 255707 (45.4%) | 361 (40.6%) | 256068 (45.4%) |
| neutrophils | 240693 (42.8%) | 333 (37.5%) | 241026 (42.8%) |
| glomerular filtration rate | 163057 (29.0%) | 215 (24.2%) | 163272 (29.0%) |
| platelet mean volume | 373788 (66.4%) | 572 (64.3%) | 374360 (66.4%) |

### 4.2 Predictive performance

Table 6 shows the AUC and PPV for models trained on patients aged 18 and above, setting the false positive rate at 0.02 for PPV evaluation. The ensemble model improved the AUC (0.801) by 0.03 (95% confidence interval 0.02-0.04) compared to the baseline (0.773) and showed consistent superiority and less variability in AUC over time (0.776-0.817). It also significantly enhanced the PPV, both overall (0.0173) and across all periods (0.0119-0.032). The improvement in overall PPV was estimated to be 0.0057 (95% confidence interval 0.0022 - 0.0092). Similar patterns were observed in patients aged 40 and above, although their AUCs were generally lower due to the reduced predictive power of age in this older subgroup. For detailed results, see the Supplementary Material (Table A5). The ensemble model trained using all-stage NSCLC cases and using only early-stage NSCLC cases had very similar performance. In the remainder of this section, we report performance based on the early-stage ensemble model.

**Table 6:** Area under the receiver operating characteristic curve (AUC) and Positive Predictive Value (PPV) for predicting early-stage NSCLC diagnosis, both overall and by 3-year period among patients aged 18 and above: comparing the baseline model and the ensemble model.

|  | **Overall** | **2007-**<br>**2009** | **2010-**<br>**2012** | **2013-**<br>**2015** | **2016-**<br>**2018** | **2019-**<br>**2021** |
| --- | --- | --- | --- | --- | --- | --- |
| **AUC baseline** | 0.773 | 0.801 | 0.740 | 0.769 | 0.794 | 0.746 |
| **AUC ensemble** | 0.801 | 0.817 | 0.785 | 0.798 | 0.816 | 0.776 |
| **AUC ensemble (early-stage)** | 0.801 | 0.815 | 0.777 | 0.796 | 0.818 | 0.781 |
| **incidence** | 0.0018 | 0.0011 | 0.0014 | 0.0021 | 0.0024 | 0.0015 |
| **PPV baseline** | 0.0116 | 0.0075 | 0.0067 | 0.0171 | 0.0111 | 0.0091 |
| **PPV ensemble** | 0.0173 | 0.0148 | 0.0132 | 0.0321 | 0.0171 | 0.0119 |
| **PPV ensemble (early-stage)** | 0.0173 | 0.0175 | 0.0136 | 0.0261 | 0.0165 | 0.0125 |

Table 7 shows the predictive performance of our ensemble model across patient subgroups, including the non-white population and by gender. Current lung cancer screening guidelines recommend screening for individuals aged 50 to 80 years with a 20-pack-year smoking history, who are either current smokers or have quit within the past 15 years (8). We studied the performance among non-smokers, a group not addressed by existing guidelines. Smoker status was determined using health survey data available for some MGB Biobank patients. We observed that the ensemble model outperformed the baseline model in all subgroups considered in terms of AUC and PPV; the ensemble model showed substantial improvements over the baseline model among non-smokers, which is a subgroup not well captured by the current screening guidelines; the ensemble model showed significant improvements over the baseline model among heavy-smokers (≥ 20 pack-year); and the ensemble model demonstrated superior performance among non-white patients.

**Table 7:** Area under the receiver operating characteristic curve (AUC) and Positive Predictive Value (PPV) for predicting early-stage NSCLC diagnosis among subgroups of patients, comparing the baseline model and the ensemble model.

|  | AUC |  | PPV |  | Incidence of early-stage NSCLC |
| --- | --- | --- | --- | --- | --- |
| subgroup | ensemble | baseline | ensemble | baseline |  |
| **females** | 0.820 | 0.789 | 0.0182 | 0.0123 | 0.0016 |
| **males** | 0.776 | 0.752 | 0.0159 | 0.0064 | 0.0019 |
| **white** | 0.791 | 0.765 | 0.0178 | 0.0123 | 0.0018 |
| **non-white** | 0.850 | 0.806 | 0.0078 | 0.0060 | 0.0009 |
| **smokers ($\geq 20$ pack-year)\*** | 0.737 | 0.670 | 0.0502 | 0.0135 | 0.0063 |
| **non-smokers** | 0.768 | 0.696 | 0.0072 | 0.0057 | 0.0006 |
| **not screened for lung cancer in the year prior to diagnosis<sup>†</sup>** | 0.799 | 0.771 | 0.0196 | 0.0118 | 0.0017 |

\*including both current and past smokers; † defined as not having any procedure codes corresponding to lung cancer screening via low-dose CT in the calendar year prior to the first lung cancer diagnosis.

Lastly, we extracted codified data related to lung cancer screening via low-dose CT and examined the model performance among patients not screened for lung cancer. The proportion of patients in the MGB Biobank who underwent lung cancer screening was generally very low (<0.7% in every yearly cohort), and only around 6% of the NSCLC cases had undergone lung cancer screening in the year prior to their first lung cancer diagnosis.

The last row in Table 7 shows the performance of the ensemble and baseline models among patients aged 18 and above who were not screened for lung cancer. The ensemble model again showed significant improvements over the baseline model. In fact, the performance both overall and by 3-year periods are very similar to those in Table 6 in the entire Biobank cohort, due to the low number of patients screened for lung cancer.

### 4.3 Feature importance

Besides patient demographics (age, sex, race), social determinants of health (area deprivation index and social vulnerability index), and healthcare utilization, we identified 117 EHR-derived features predictive for early NSCLC detection using early-stage patients, and 127 EHR-derived features with all-stage patients. Figure 4 shows the feature importance measured by the Shapley value (35) of the top 30 EHR-derived features comparing early-stage to all-stage models. Highly predictive features included smoking, socioeconomic conditions, relevant lab test results, and chronic lung diseases. Although the ranking of individual features differed slightly between the all-stage and early-stage models, the set of important features identified were very similar. This also explained the very similar predictive performance of these two models.

**Figure 4:**
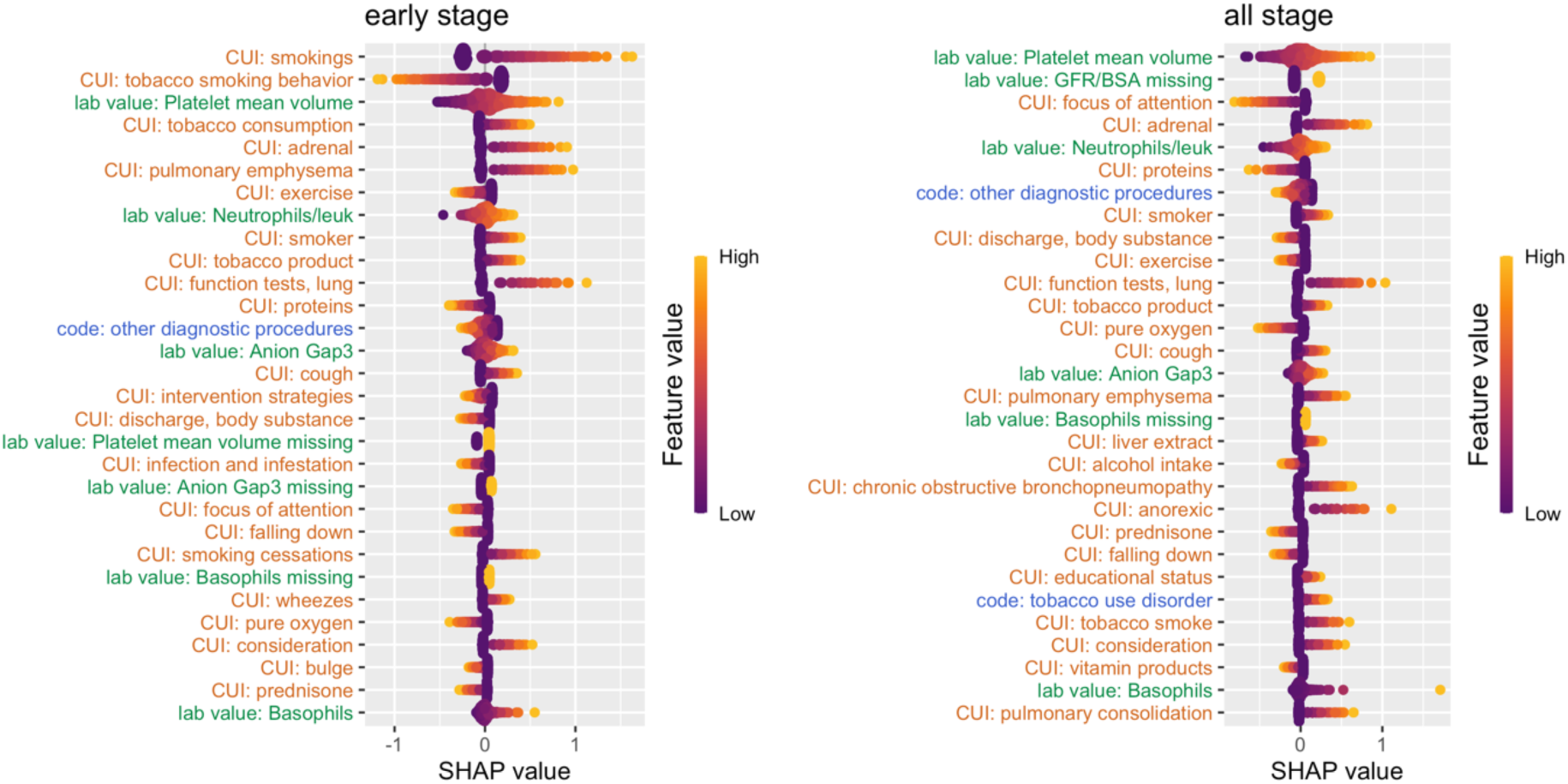
Feature importance for Predicting 1-Year Risk of early-stage Non-Small Cell Lung Cancer (NSCLC). Feature importance is measured by average absolute Shapley value. Top 30 EHR-derived features are presented with decreasing feature importance from top to bottom. A positive Shapley value indicates an increased risk of lung cancer diagnosis. Color of text on the y-axis indicates feature category: Green (Numerical Lab Values or missing indicators of numerical lab values, also indicated by “lab value”), Orange (Concept Unique Identifiers Extracted from Natural Language Processing, also indicated by “CUI”), Blue (Codified Features, also indicated by “code”).

## 5 DISCUSSION AND CONCLUSION

Effectively leveraging two EHR cohorts extracted from the MGB EHR databases, we identified EHR-derived features associated with early NSCLC diagnosis. Consequently, we developed 1- year risk prediction models for early NSCLC with notably high predictive performance. Specifically, the models achieved an AUC of up to 0.801 for the patient population aged 18 and above, and an AUC of 0.757 for patients aged 40 and above, in predicting early-stage NSCLC diagnosis. Our prediction model presents a different approach compared to existing EHR-based lung cancer prediction models. Firstly, it was evaluated prospectively, offering enhanced reliability and applicability in real-world scenarios. Secondly, when evaluating the model performance, we focused on predicting early-stage diagnosis in particular, which aligns with our goal of early cancer detection. Additionally, our model was constructed using features that do not directly involve lung cancer diagnosis, thereby avoiding reliance on features like CT scans and nodule findings.

Our model identified known risk factors such as older age and socioeconomic factors, tobacco use and chronic obstructive pulmonary diseases (COPD). Other important EHR-derived risk predictors included signs and symptoms associated with the respiratory system including coughing, and relevant lab test results including platelet mean volume and number of neutrophils in blood. Notably, these lab tests were also found to be predictive of NSCLC in prior EHR-based studies (17).

We found that lung cancer screening rates were low, as shown by EHR data, with only a small fraction of diagnosed patients having been screened the year before their diagnosis. This suggests a need to promote screening uptake among eligible patients within the MGB healthcare system and to enhance current guidelines with tools that identify high-risk individuals early. Our study underscored low screening rates and demonstrated that our prediction model performs well in subpopulations not adequately covered by existing screening recommendations, highlighting the potential of an EHR-based prediction tool in clinical settings to support screening efforts. However, the lack of smoking pack-year data and difficulties in extracting such information from clinical notes limited our ability to compare our model’s high-risk population identification with those of current screening guidelines or established models like the PLCO^m^2012 (13–15).

We found the proportion of lung cancer cases diagnosed at the metastatic stage was consistent between the LC mart and the MGB Biobank data, which is significantly lower than that reported in the general population (2). The higher proportion of early-stage patients in our data enabled us to train and evaluate an ensemble learning model using early-stage NSCLC patients only. The exact reason behind the difference in the stage distribution between the MGB EHR population and the general population is unclear. It is worth investigating in future studies how this population shift impacts the transferability of our model outside of the MGB system.

## Data Availability

Patient-level data are protected due to privacy concerns, and all study results including model parameters in the present study are available upon reasonable request to the authors.

## Footnotes

1 A major update to the MGB EHR database occurred in 2006. EHR data within the four years prior to the first lung cancer diagnosis were used in the self-control design, while data in the year prior to the first lung cancer diagnosis were used in the case-control design. This inclusion criterion ensured that all data used in the modeling were after 2006 and at the same time maximized the number of patients included.

## REFERENCES

1. Siegel RL, Giaquinto AN, Jemal A. Cancer Statistics, 2024. CA Cancer J Clin 2024;74(1):12-49.

2. Surveillance, Epidemiology, and End Results Program, National Cancer Institute. Cancer Stat Facts. [accessed June 2024]. Available from: https://seer.cancer.gov/statfacts/.

3. Lancet T. Late-Stage Cancer Detection in the USA Is Costing Lives. 2010. Lancet 376;9756:1873.

4. Raoof S, Lee RJ, Jajoo K, Mancias JD, Rebbeck TR, Skates SJ. Multicancer early detection technologies: a review informed by past cancer screening studies. Cancer Epidemiol Biomarkers Prev 2022;31(6):1139–1145.

5. Walter FM, Rubin G, Bankhead C, Morris HC, Hall N, Mills K, et al. Symptoms and other factors associated with time to diagnosis and stage of lung cancer: a prospective cohort study. Br J Cancer 2015;112 Suppl 1(S1):S6-13.

6. Ruano-Raviña A, Provencio M, Calvo de Juan V, Carcereny E, Moran T, Rodriguez-Abreau D, et al. Lung cancer symptoms at diagnosis: results of a nationwide registry study. ESMO Open. 2020;5(6):e001021.

7. Smith RA, Andrews KS, Brooks D, Fedewa SA, Manassaram-Baptiste D, Saslow D, et al. Cancer screening in the United States, 2019: A review of current American Cancer Society guidelines and current issues in cancer screening. CA Cancer J Clin 2019;69(3):184-210.

8. Krist AH, Davidson KW, Mangione CM, Barry MJ, Cabana M, Caughey AB et al. Screening for lung cancer: US Preventive Services Task Force recommendation statement. JAMA 2021;325(10):962–970.

9. Jemal A, Fedewa SA. Lung Cancer Screening With Low-Dose Computed Tomography in the United States—2010 to 2015. JAMA Oncol 2017;3(9):1278–81.

10. American Cancer Society. Cancer Facts & Figures 2024. 2024 [accessed June 2024]. Available from: https://www.cancer.org/research/cancer-facts-statistics/all-cancer-facts-figures/2024-cancer-facts-figures.html

11. Thandra KC, Barsouk A, Saginala K, Aluru JS, Barsouk A. Epidemiology of Lung Cancer. Contemp Oncol 2021;25(1):45–52.

12. Gildea TR, DaCosta Byfield S, Hogarth DK, Wilson DS, Quinn CC. A Retrospective Analysis of Delays in the Diagnosis of Lung Cancer and Associated Costs. Clinicoecon Outcomes Res 2017;9:261–69.

13. Tammemägi MC, Pinsky PF, Caporaso NE, Kvale PA, Hocking WG, Church TR, et al. Lung cancer risk prediction: prostate, lung, colorectal and ovarian cancer screening trial models and validation. J Natl Cancer Inst 2011;103(13):1058–1068.

14. Tammemägi MC, Katki HA, Hocking WG, Church TR, Caporaso NE, Kvale PA, et al. Selection criteria for lung-cancer screening. N Engl J Med 2013;368(8):728–736.

15. Katki HA, Kovalchik SA, Petito LC, Cheung LC, Jacobs E, Jemal A, et al. Implications of nine risk prediction models for selecting ever-smokers for computed tomography lung cancer screening. Ann Intern Med 2018;169(1):10–19.

16. Muller DC, Johansson M, Brennan P. (2017). Lung cancer risk prediction model incorporating lung function: development and validation in the UK Biobank prospective cohort study. J Clin Oncol 2017;35(8):861-869.

17. Gould MK, Huang BZ, Tammemagi MC, Kinar Y, Shiff R. Machine Learning for Early Lung Cancer Identification Using Routine Clinical and Laboratory Data. Am J Respir Crit Care Med 2021;204(4):445–53.

18. Wang X, Zhang Y, Hao S, Zheng L, Liao J, Ye C, Xia M, et al. Prediction of the 1- Year Risk of Incident Lung Cancer: Prospective Study Using Electronic Health Records from the State of Maine. J Med Internet Res 2019;21(5):e13260.

19. Chandran U, Reps J, Yang R, Vachani A, Maldonado F, Kalsekar I. Machine learning and real-world data to predict lung cancer risk in routine care. Cancer Epidemiol Biomarkers Prev 2023;32(3):337–343.

20. Yuan Q, Cai T, Hong C, Du M, Johnson BE, Lanuti M, et al. 2021. Performance of a Machine Learning Algorithm Using Electronic Health Record Data to Identify and Estimate Survival in a Longitudinal Cohort of Patients With Lung Cancer. JAMA Netw Open 2021;4(7):e2114723.

21. Castro VM, Gainer V, Wattanasin N, Benoit B, Cagan A, Ghosh B, et al. The Mass General Brigham Biobank Portal: an i2b2-based data repository linking disparate and high-dimensional patient data to support multimodal analytics. J Am Med Inform Assoc 2022;29(4):643–651.

22. Yu S, Cai T, Cai T. NILE: fast natural language processing for electronic health records. 2013. arXiv preprint arXiv:1311.6063.

23. Bodenreider O. The unified medical language system (UMLS): integrating biomedical terminology. Nucleic Acids Res 2004;32(suppl_1):D267-D270.

24. Centers for Disease Control and Prevention. Agency for Toxic Substances and Disease Registry. CDC/ATSDR social vulnerability index 2020. Available from: https://www.atsdr.cdc.gov/placeandhealth/svi/index.html.

25. University of Wisconsin Health Innovation Program. HIPxChange: sharing to transform healthcare. Area Deprivation Index. 2012. Available from: http://www.hipxchange.org/ADI.

26. Li X, Yuan EY, Duan R, Cai T. Early Detection of Diseases in Electronic Health Records: A Comparative Approach Combining Three Study Designs. In Preparation. Published online 2024+.

27. Bastarache L. Using phecodes for research with the electronic health record: from PheWAS to PheRS. Annu Rev Biomed Data Sci 2021;4(1):1–19.

28. Elixhauser AS, Palmer L. Clinical Classifications Software (CCS): Agency for Healthcare Research and Quality 2014. Available from: http://www.hcup-us.ahrq.gov/toolssoftware/ccs/ccs.jsp.

29. Liu S, Ma W, Moore R, Ganesan V, Nelson S. RxNorm: prescription for electronic drug information exchange. IT Prof 2005;7(5):17–23.

30. Huff SM, Rocha RA, McDonald CJ, De Moor GJ, Fiers T, Bidgood Jr WD, et al. Development of the logical observation identifier names and codes (LOINC) vocabulary. J Am Med Inform Assoc 1998;5(3):276–292.

31. Xiong X, Sweet SM, Liu M, Hong C, Bonzel CL, Ayakulangara Panickan V, et al. Knowledge-Driven Online Multimodal Automated Phenotyping System. medRxiv 2023:2023.09.

32. Benjamini Y, Hochberg Y. Controlling the false discovery rate: a practical and powerful approach to multiple testing. J R Stat Soc Ser B Methodol 1995;57(1):289–300.

33. Van Buuren S, Groothuis-Oudshoorn K. (2011). MICE: Multivariate imputation by chained equations in R. J Stat Softw 2011;*45*:1–67.

34. Pinsky P. Electronic health records and machine learning for early detection of lung cancer and other conditions: thinking about the path ahead. Am J Respir Crit Care Med 2021;204(4):389–390.

35. Štrumbelj E, Igor K. Explaining prediction models and individual predictions with feature contributions. Knowl Inf Syst 2014;41:647–665.

