## Supplementary Material for "Early detection of non-small cell lung cancer using electronic health record data"

Appendix I – Supplemental Tables, Figures and Graphs

In this Appendix, we present additional tables on the patient characteristics for all patients in the Lung Cancer (LC) mart, as well as the patients included in the self-controlled design, case-control design, and the yearly cohorts in the prospective modeling stage.

**Table A1:** Demographics and clinical attributes of the lung cancer cohort, overall and stratified by stage (stage 1-3 versus stage 4). For continuous variables we present mean (standard deviation), and for categorical variables we present count (percentage).

| | Stage I-III (N=27816) | Stage IV (N=11383) | Overall (N=63915) |
|---|---|---|---|
| **sex** | | | |
| female | 15137 (54.4%) | 6019 (52.9%) | 33335 (52.2%) |
| male | 12678 (45.6%) | 5362 (47.1%) | 30565 (47.8%) |
| unknown | 1 (0.0%) | 2 (0.0%) | 15 (0.0%) |
| **age** | | | |
| mean (SD) | 66.3 (11.1) | 63.6 (11.7) | 65.4 (12.0) |
| **race** | | | |
| black | 747 (2.7%) | 374 (3.3%) | 1689 (2.6%) |
| other | 1150 (4.1%) | 772 (6.8%) | 2846 (4.5%) |
| unknown | 1295 (4.7%) | 642 (5.6%) | 6402 (10.0%) |
| white | 24624 (88.5%) | 9595 (84.3%) | 52978 (82.9%) |
| **first diagnosis year** | | | |
| before 1994 | 138 (0.5%) | 27 (0.2%) | 558 (0.9%) |
| 1994-1997 | 1509 (5.4%) | 297 (2.6%) | 4560 (7.1%) |
| 1998-2001 | 2399 (8.6%) | 694 (6.1%) | 7025 (11.0%) |
| 2002-2005 | 3521 (12.7%) | 1272 (11.2%) | 8676 (13.6%) |
| 2006-2009 | 4831 (17.4%) | 1817 (16.0%) | 10209 (16.0%) |
| 2010-2013 | 5224 (18.8%) | 2424 (21.3%) | 11049 (17.3%) |
| 2014-2017 | 6219 (22.4%) | 3088 (27.1%) | 14021 (21.9%) |
| 2018-2021 | 3975 (14.3%) | 1764 (15.5%) | 7817 (12.2%) |
| **months of prior EHR records** | | | |

| | | | |
|---|---|---|---|
| mean (SD) | 112 (98.4) | 79.8 (88.7) | 79.1 (92.3) |
| median [Min, Max] | 88.0 [0, 737] | 39.0 [0, 669] | 37.0 [0, 737] |
| **Histology** | | | |
| non-small cell | 26254 (94.4%) | 9829 (86.3%) | 46647 (73.0%) |
| adenocarcinoma | 12398 (44.6%) | 5123 (45.0%) | 22251 (34.8%) |
| squamous cell cancer | 4612 (16.6%) | 932 (8.2%) | 7149 (11.2%) |
| large cell cancer | 203 (0.7%) | 77 (0.7%) | 421 (0.7%) |
| small cell | 1562 (5.6%) | 1554 (13.7%) | 5368 (8.4%) |
| missing | 0 (0%) | 0 (0%) | 11900 (18.6%) |

**Table A2:** Age, smoking, comorbidities, and lab values comparing cancer-diagnosis and control windows among NSCLC patients who met all inclusion criteria. Smoking is defined as the occurrence of smoking-related code and/or CUI; comorbidity is defined as the occurrence of relevant diagnosis codes.

| | control period (N=3546) | cancer diagnosis period (N=3546) | Overall |
|---|---|---|---|
| **comorbidities** | | | |
| myocardial infarction | 119 (3.4%) | 128 (3.6%) | 247 (3.5%) |
| congestive heart failure | 207 (5.8%) | 293 (8.3%) | 500 (7.1%) |
| peripheral vascular disease | 303 (8.5%) | 389 (11.0%) | 692 (9.8%) |
| cerebrovascular disease | 229 (6.5%) | 273 (7.7%) | 502 (7.1%) |
| dementia | 27 (0.8%) | 62 (1.7%) | 89 (1.3%) |
| chronic pulmonary disease | 528 (14.9%) | 742 (20.9%) | 1270 (17.9%) |
| rheumatic disease | 97 (2.7%) | 115 (3.2%) | 212 (3.0%) |
| peptic ulcer disease | 20 (0.6%) | 36 (1.0%) | 56 (0.8%) |
| liver disease | 110 (3.1%) | 142 (4.0%) | 252 (3.6%) |
| diabetes | 368 (10.4%) | 416 (11.7%) | 784 (11.1%) |
| hemiplegia or paraplegia | 17 (0.5%) | 20 (0.6%) | 37 (0.5%) |

| | | | |
|---|---|---|---|
| renal disease | 143 (4.0%) | 218 (6.1%) | 361 (5.1%) |
| HIV/AIDS | 0 (0%) | 0 (0%) | 0 (0%) |
| **smoking** | | | |
| smoker | 1261 (35.6%) | 1887 (53.2%) | 3148 (44.4%) |
| **healthcare utilization** | | | |
| mean (SD) | 1.78 (1.37) | 2.19 (1.59) | 1.99 (1.50) |
| **lab values** | | | |
| monocytes (%) | 7.27 (1.89) | 7.69 (2.03) | 7.48 (1.97) |
| anion gap 3(mmol/L) | 11.8 (2.31) | 12.3 (2.50) | 12.0 (2.42) |
| basophils (k/mm^3) | 0.0405 (0.0228) | 0.0413 (0.0224) | 0.0409 (0.0226) |
| neutrophils (%) | 63.8 (7.62) | 64.4 (8.08) | 64.1 (7.86) |
| glomerular filtration rate (ml/min/1.73m^2) | 61.5 (9.81) | 61.8 (12.2) | 61.6 (11.1) |
| platelet mean volume (fl) | 10.4 (0.488) | 10.4 (0.635) | 10.4 (0.566) |
| **lab value missing*** | | | |
| monocytes | 2435 (68.7%) | 2003 (56.5%) | 4438 (62.6%) |
| anion gap 3 | 2297 (64.8%) | 1707 (48.1%) | 4004 (56.5%) |
| basophils | 2521 (71.1%) | 2048 (57.8%) | 4569 (64.4%) |
| neutrophils | 2443 (68.9%) | 2011 (56.7%) | 4454 (62.8%) |
| glomerular filtration rate | 2175 (61.3%) | 1624 (45.8%) | 3799 (53.6%) |
| platelet mean volume | 3322 (93.7%) | 2524 (71.2%) | 5846 (82.4%) |

* Lab value is treated as missing if the lab test was not done or if there is evidence that the test was done but the result is missing in the EHR. In either case, the missing lab value was imputed through multivariate imputation by chained equations.

**Table A3:** Baseline characteristics of all patients in the case-control design. Cases are NSCLC patients from the lung cancer cohort; and controls are patients free of lung cancer in the MBG Biobank, matched based on calendar year, healthcare utilization, and patient demographics including age, sex, and race.

| | control (N=12686) | case (N=6354) | overall (N=19040) |
|---|---|---|---|
| **age** | | | |

| | | | |
|---|---|---|---|
| mean (SD) | 67.2 (11.6) | 67.6 (10.7) | 67.4 (11.3) |
| **sex** | | | |
| female | 7293 (57.5%) | 3702 (58.3%) | 10995 (57.7%) |
| male | 5393 (42.5%) | 2652 (41.7%) | 8045 (42.3%) |
| **race** | | | |
| black | 417 (3.3%) | 253 (4.0%) | 670 (3.5%) |
| other | 616 (4.9%) | 331 (5.2%) | 947 (5.0%) |
| unknown | 206 (1.6%) | 149 (2.3%) | 355 (1.9%) |
| white | 11447 (90.2%) | 5621 (88.5%) | 17068 (89.6%) |
| **Calendar year of follow up** | | | |
| 2006-2008 | 1992 (15.7%) | 993 (15.6%) | 2985 (15.7%) |
| 2009-2011 | 2238 (17.6%) | 1194 (18.8%) | 3432 (18.0%) |
| 2012-2014 | 2691 (21.2%) | 1318 (20.7%) | 4009 (21.1%) |
| 2015-2017 | 3063 (24.1%) | 1650 (26.0%) | 4713 (24.8%) |
| 2018-2020 | 2702 (21.3%) | 1199 (18.9%) | 3901 (20.5%) |
| **smoking** | | | |
| smoker | 3325 (26.2%) | 3442 (54.2%) | 6767 (35.5%) |
| **healthcare utilization** | | | |
| mean (SD) | 2.33 (1.15) | 2.29 (1.37) | 2.32 (1.23) |
| **comorbidities** | | | |
| myocardial infarction | 438 (3.5%) | 242 (3.8%) | 680 (3.6%) |
| congestive heart failure | 1105 (8.7%) | 487 (7.7%) | 1592 (8.4%) |
| peripheral vascular disease | 896 (7.1%) | 612 (9.6%) | 1508 (7.9%) |
| cerebrovascular disease | 1106 (8.7%) | 461 (7.3%) | 1567 (8.2%) |
| dementia | 355 (2.8%) | 123 (1.9%) | 478 (2.5%) |
| chronic pulmonary disease | 549 (4.3%) | 898 (14.1%) | 1447 (7.6%) |
| rheumatic disease | 581 (4.6%) | 174 (2.7%) | 755 (4.0%) |
| peptic ulcer disease | 75 (0.6%) | 54 (0.8%) | 129 (0.7%) |
| liver disease | 532 (4.2%) | 295 (4.6%) | 827 (4.3%) |
| diabetes | 1518 (12.0%) | 754 (11.9%) | 2272 (11.9%) |
| hemiplegia or paraplegia | 117 (0.9%) | 41 (0.6%) | 158 (0.8%) |

| | | | |
|---|---|---|---|
| renal disease | 754 (5.9%) | 354 (5.6%) | 1108 (5.8%) |
| HIV/AIDS | 0 (0%) | 0 (0%) | 0 (0%) |
| **lab values** | | | |
| monocytes (%) | 7.28 (2.25) | 7.41 (2.09) | 7.32 (2.21) |
| anion gap 3(mmol/L) | 11.0 (2.91) | 12.0 (2.58) | 11.3 (2.85) |
| basophils (k/mm^3) | 0 (0) | 0.0411 (0.0215) | 0.0123 (0.0222) |
| neutrophils (%) | 61.4 (9.36) | 65.1 (8.47) | 62.5 (9.26) |
| glomerular filtration rate (ml/min/1.73m^2) | 64.5 (14.3) | 62.4 (12.8) | 63.9 (13.9) |
| platelet mean volume (fl) | 9.89 (0.723) | 10.4 (0.615) | 10.0 (0.731) |
| **lab missing** | | | |
| monocytes | 6194 (48.8%) | 3251 (51.2%) | 9445 (49.6%) |
| anion gap 3 | 5196 (41.0%) | 2941 (46.3%) | 8137 (42.7%) |
| basophils | 6556 (51.7%) | 3374 (53.1%) | 9930 (52.2%) |
| neutrophils | 6241 (49.2%) | 3249 (51.1%) | 9490 (49.8%) |
| glomerular filtration rate | 4359 (34.4%) | 2731 (43.0%) | 7090 (37.2%) |
| platelet mean volume | 9460 (74.6%) | 4820 (75.9%) | 14280 (75.0%) |

**Table A4**: Patient demographics, smoking status, comorbidities, and lab tests in the MGB Biobank patients in the prospective modeling stage, comparing NSCLC cases versus NSCLC-free patients.

| | NSCLC-free (N=562637) | NSCLC case (N=1180) | Overall (N=563817) |
|---|---|---|---|
| **age** | | | |
| mean (SD) | 50.4 (15.3) | 64.2 (12.0) | 50.4 (15.3) |
| **race** | | | |
| black | 32865 (5.8%) | 37 (3.1%) | 32902 (5.8%) |
| other | 40900 (7.3%) | 37 (3.1%) | 40937 (7.3%) |
| unknown | 14674 (2.6%) | 26 (2.2%) | 14700 (2.6%) |
| white | 474198 (84.3%) | 1080 (91.5%) | 475278 (84.3%) |
| **sex** | | | |

| | | | |
|---|---|---|---|
| female | 326615 (58.1%) | 635 (53.8%) | 327250 (58.0%) |
| male | 236006 (41.9%) | 545 (46.2%) | 236551 (42.0%) |
| unknown | 16 (0.0%) | 0 (0%) | 16 (0.0%) |
| **area deprivation index (national)** | | | |
| mean (SD) | 13.2 (12.6) | 15.0 (14.8) | 13.2 (12.6) |
| **area deprivation index (state)** | | | |
| mean (SD) | 3.73 (2.36) | 3.97 (2.52) | 3.73 (2.36) |
| **social vulnerability index** | | | |
| mean (SD) | 0.375 (0.198) | 0.370 (0.202) | 0.375 (0.198) |
| **calendar year of follow-up period** | | | |
| 2007-2009 | 74585 (13.3%) | 85 (7.2%) | 74670 (13.2%) |
| 2010-2012 | 90964 (16.2%) | 138 (11.7%) | 91102 (16.2%) |
| 2013-2015 | 112276 (20.0%) | 248 (21.0%) | 112524 (20.0%) |
| 2016-2018 | 136871 (24.3%) | 419 (35.5%) | 137290 (24.4%) |
| 2019-2021 | 147941 (26.3%) | 290 (24.6%) | 148231 (26.3%) |
| **healthcare utilization** | | | |
| mean (SD) | 2.55 (1.23) | 2.93 (1.45) | 2.55 (1.23) |
| **comorbidities** | | | |
| myocardial infarction | 11932 (2.1%) | 62 (5.3%) | 11994 (2.1%) |
| congestive heart failure | 33657 (6.0%) | 151 (12.8%) | 33808 (6.0%) |
| peripheral vascular disease | 25806 (4.6%) | 122 (10.3%) | 25928 (4.6%) |
| cerebrovascular disease | 25027 (4.4%) | 109 (9.2%) | 25136 (4.5%) |
| dementia | 6654 (1.2%) | 27 (2.3%) | 6681 (1.2%) |
| chronic pulmonary disease | 63914 (11.4%) | 353 (29.9%) | 64267 (11.4%) |
| rheumatic disease | 26905 (4.8%) | 75 (6.4%) | 26980 (4.8%) |
| peptic ulcer disease | 3217 (0.6%) | 10 (0.8%) | 3227 (0.6%) |
| liver disease | 36806 (6.5%) | 111 (9.4%) | 36917 (6.5%) |
| diabetes | 64686 (11.5%) | 220 (18.6%) | 64906 (11.5%) |
| hemiplegia or paraplegia | 4473 (0.8%) | 15 (1.3%) | 4488 (0.8%) |

| | | | |
|---|---|---|---|
| renal disease | 27927 (5.0%) | 140 (11.9%) | 28067 (5.0%) |
| HIV/AIDS | 2730 (0.5%) | 4 (0.3%) | 2734 (0.5%) |
| **smoking** | | | |
| smoker | 158453 (28.2%) | 625 (53.0%) | 159078 (28.2%) |
| **lab value** | | | |
| monocytes (%) | 7.37 (2.22) | 7.78 (2.45) | 7.37 (2.22) |
| anion gap 3(mmol/L) | 11.6 (2.86) | 12.0 (2.91) | 11.6 (2.86) |
| basophils (k/mm^3) | 0.0410 (0.0262) | 0.0441 (0.0261) | 0.0410 (0.0262) |
| neutrophils (%) | 61.3 (9.69) | 63.3 (9.94) | 61.3 (9.70) |
| glomerular filtration rate (ml/min/1.73m^2) | 70.3 (18.0) | 63.7 (17.2) | 70.3 (18.0) |
| platelet mean volume (fl) | 10.4 (0.699) | 10.4 (0.757) | 10.4 (0.699) |
| **lab missing** | | | |
| monocytes | 239650 (42.6%) | 433 (36.7%) | 240083 (42.6%) |
| anion gap 3 | 200981 (35.7%) | 369 (31.3%) | 201350 (35.7%) |
| basophils | 255707 (45.4%) | 469 (39.7%) | 256176 (45.4%) |
| neutrophils | 240693 (42.8%) | 434 (36.8%) | 241127 (42.8%) |
| glomerular filtration rate | 163057 (29.0%) | 273 (23.1%) | 163330 (29.0%) |
| platelet mean volume | 373788 (66.4%) | 715 (60.6%) | 374503 (66.4%) |

Appendix II – Predictive performance of ensemble model among patients aged 40 and above

In Table A5, we present the predictive performance comparing the ensemble model trained using our three-stage design and the baseline model that only uses demographics and smoking information, among patients aged 40 and above. We observe that the ensemble model consistently outperforms the baseline model in terms of the area under the receiver operating characteristic curve (AUC), both overall and by 3-year periods. The ensemble model also significantly improves the Positive Predictive Value (PPV) compared with the baseline model.

**Table A5**: Area under the receiver operating characteristic curve (AUC) and Positive Predictive Value (PPV) for predicting early-stage NSCLC diagnosis, both overall and by 3-year period among patients aged 40 and above: comparing the baseline model and the ensemble model.

| | Overall | 2007-2009 | 2010-2012 | 2013-2015 | 2016-2018 | 2019-2021 |
|---|---|---|---|---|---|---|
| **AUC baseline** | 0.722 | 0.737 | 0.725 | 0.694 | 0.751 | 0.693 |
| **AUC ensemble** | 0.754 | 0.754 | 0.773 | 0.730 | 0.771 | 0.733 |
| **AUC ensemble (early-stage)** | 0.757 | 0.750 | 0.764 | 0.731 | 0.778 | 0.746 |
| **incidence** | 0.0022 | 0.0014 | 0.0017 | 0.0027 | 0.0031 | 0.0018 |
| **PPV baseline** | 0.0127 | 0.0128 | 0.0043 | 0.0187 | 0.0094 | 0.0102 |
| **PPV ensemble** | 0.0173 | 0.0199 | 0.0184 | 0.0265 | 0.0234 | 0.0117 |
| **PPV ensemble (early-stage)** | 0.0196 | 0.0213 | 0.0153 | 0.0337 | 0.0188 | 0.0122 |

Appendix III — Comparing different model specifications in terms of predictive performance in the prospective modeling stage

We compare the predictive performance in detecting early-stage NSCLC among patients aged 18 and above, varying the following aspects in training and calibrating the risk prediction models in the self-controlled and case-control design: (1) using early-stage patients or patients of all stage (stage); (2) whether we restrict to the subset of features in the self-controlled design that also pass the marginal screening in the case-control design (intersect SC with CC); and (3) whether we restrict to the subset of features in the case-control design that also pass the marginal screening in the self-controlled design (intersect CC with SC). Here, "baseline" stands for the baseline model that only includes patient demographics and smoking; and "ensemble" stands for the model trained using our proposed ensemble approach. We observe that different model specifications result in very similar performance overall and significant improvements over the baseline model. Similar results were observed when we restrict to patients aged 40 and above.

**Table A6**: Predictive performance in detecting early-stage NSCLC among patients aged 18 or above: a comparison among different model specifications in training and calibration.

| stage | Intersect SC with CC | Intersect CC with SC | Baseline AUC | Ensemble AUC | Baseline PPV | Ensemble PPV |
|---|---|---|---|---|---|---|
| Early | No | No | 0.773 | 0.796 | 0.0116 | 0.0183 |
| Early | No | Yes | 0.773 | 0.799 | 0.0116 | 0.0154 |
| Early | Yes | No | 0.773 | 0.799 | 0.0116 | 0.0202 |
| Early | Yes | Yes | 0.773 | 0.801 | 0.0116 | 0.0173 |
| All | No | No | 0.774 | 0.797 | 0.0106 | 0.0162 |
| All | No | Yes | 0.774 | 0.798 | 0.0106 | 0.0181 |
| All | Yes | No | 0.774 | 0.798 | 0.0106 | 0.0203 |
| All | Yes | Yes | 0.774 | 0.801 | 0.0106 | 0.0173 |
